# Development of a Rapid Point-Of-Care Test that Measures Neutralizing Antibodies to SARS-CoV-2

**DOI:** 10.1101/2020.12.15.20248264

**Authors:** Douglas F. Lake, Alexa J. Roeder, Erin Kaleta, Paniz Jasbi, Kirsten Pfeffer, Calvin Koelbel, Sivakumar Periasamy, Natalia Kuzmina, Alexander Bukreyev, Thomas E. Grys, Liang Wu, John R Mills, Kathrine McAulay, Maria Gonzalez-Moa, Alim Seit-Nebi, Sergei Svarovsky

**Affiliations:** School of Life Sciences, Arizona State University, Tempe AZ, USA; Mayo Clinic Arizona, Department of Laboratory Medicine and Pathology, Scottsdale, AZ, USA; College of Health Solutions, Arizona State University, Phoenix AZ, USA; Department of Pathology, University of Texas Medical Branch at Galveston, Galveston, TX USA; Galveston National Laboratory, University of Texas Medical Branch at Galveston, Galveston, TX USA; Department of Microbiology and Immunology University of Texas Medical Branch at Galveston, Galveston, TX USA; Mayo Clinic Rochester, Department of Laboratory Medicine and Pathology, Rochester, MN USA; Axim Biotechnologies Inc, San Diego, CA USA

**Keywords:** Neutralizing Antibodies, COVID-19, SARS-CoV-2, Lateral Flow Assay, RBD, ACE2

## Abstract

**Background:** After receiving a COVID-19 vaccine, most recipients want to know if they are protected from infection and for how long. Since neutralizing antibodies are a correlate of protection, we developed a lateral flow assay (LFA) that measures levels of neutralizing antibodies from a drop of blood. The LFA is based on the principle that neutralizing antibodies block binding of the receptor-binding domain (RBD) to angiotensin-converting enzyme 2 (ACE2).

**Methods:** The ability of the LFA was assessed to correctly measure neutralization of sera, plasma or whole blood from patients with COVID-19 using SARS-CoV-2 microneutralization assays. We also determined if the LFA distinguished patients with seasonal respiratory viruses from patients with COVID-19. To demonstrate the usefulness of the LFA, we tested previously infected and non-infected COVID-19 vaccine recipients at baseline and after first and second vaccine doses.

**Results:** The LFA compared favorably with SARS-CoV-2 microneutralization assays with an area under the ROC curve of 98%. Sera obtained from patients with seasonal coronaviruses did not show neutralizing activity in the LFA. After a single mRNA vaccine dose, 87% of previously infected individuals demonstrated high levels of neutralizing antibodies. However, if individuals were not previously infected only 24% demonstrated high levels of neutralizing antibodies after one vaccine dose. A second dose boosted neutralizing antibody levels just 8% higher in previously infected individuals, but over 63% higher in non-infected individuals.

**Conclusions:** A rapid, semi-quantitative, highly portable and inexpensive neutralizing antibody test might be useful for monitoring rise and fall in vaccine-induced neutralizing antibodies to COVID-19.

## INTRODUCTION

Severe Acute Respiratory Syndrome Coronavirus-2 (SARS-CoV-2) causes COVID-19 and originated in Wuhan, China in December 2019 [1–3]. Vaccines continue to be tested [4,5] with the goal of preventing COVID-19 via induction of neutralizing antibodies (NAbs) and anti-viral T cells. Vaccine trials show that RNA vaccines elicit protective immunity, but durability of natural and vaccine-induced immunity is not fully known [5]. Several groups reported that up to one-third of serum samples from individuals who recovered from COVID-19 do not neutralize SARS-CoV-2 [6–8].

Whether previously infected or vaccinated, it is informative for individuals to learn if they generated high levels of NAbs so that they can resume normal activities without fear of re-infection and transmitting the virus [9–11].

Viral neutralization assays measure antibodies that block infection of host cells. The gold standard of neutralization for SARS-CoV-2 measures reduction of viral plaques or foci in microneutralization assays. These assays are slow, laborious, require highly trained personnel and a BSL3 facility. Another challenge is that neutralization assays require careful titration of virus and depend on host cells for infection, both of which add variability to the assay. These limitations prevent use of SARS-CoV-2 neutralization assays for clinical applications.

SARS-CoV-2 uses receptor binding domain (RBD) on spike protein to bind angiotensin converting enzyme 2 (ACE2) on host cells; RBD appears to be the principal neutralizing domain [12,13]. Using this knowledge, we developed a lateral flow assay (LFA) that measures levels of NAbs which block RBD from binding to ACE2. Other groups have developed RBD-ACE2-based competition ELISAs[18,19] but none have developed a rapid, highly portable, semi-quantitative test that can easily be incorporated into clinical settings or research studies where traditional laboratory or neutralization tests are not practical.

## MATERIALS AND METHODS

### Human Subjects and Samples

Serum and finger-stick blood samples were collected for this study under an Arizona State University institutional review board (IRB)-approved protocol #0601000548 and Mayo Clinic IRB protocol #20-004544. Serum samples obtained from excess clinical samples at Mayo Clinic were left over from normal workflow. COVID-19 samples ranged from 3 to 84 days post PCR positive result.

Twenty-seven control serum samples from patients with non-COVID-19 respiratory illnesses as determined by the FilmArray Respiratory Panel 2 (Biofire Diagnostics) were collected from patients from 2/14/17 – 4/6/20 as part of routine clinical workflow. All residual clinical samples were stored at 2-8°C for up to 7 days, and frozen at −80°C thereafter.

### SARS-CoV-2 Microneutralization Assay

A microneutralization assay was performed using a recombinant SARS-CoV-2 expressing mNeonGreen (SARS-CoV-2ng) as previously described [16]. Inhibitory concentrations for which 50% of virus is neutralized by serum antibodies (IC50 values) were obtained on a set of 38 COVID-19 sera. Sixty µl aliquots of SARS-CoV-2ng were pre-incubated for 1 h in 5% CO2 at 37°C with 60µl 2-fold serum dilutions in cell culture media, and 100µl were inoculated onto Vero-E6 monolayers in black polystyrene 96-well plates with clear bottoms (Corning) in duplicate. The final amount of the virus was 200 PFU/well, the starting serum dilution was 1:20 and the end dilution was 1:1280 unless an IC50 was not reached in which case serum was diluted to 1:10240. Cells were maintained in Minimal Essential Medium (ThermoFisher Scientific) supplemented by 2% FBS (HyClone) and 0.1% gentamycin in 5% CO2 at 37°C. After 2 days of incubation, fluorescence intensity of infected cells was measured at 488 nm using a Synergy 2 Cell Imaging Reader (Biotek). Signal was normalized to virus alone with no serum added and reported as percent neutralization. IC50 was calculated with GraphPad Prism 6.0 software. Work was performed in a BSL-3 biocontainment laboratory of the University of Texas Medical Branch, Galveston, TX.

### Serologic Antibody Assay

The Ortho Vitros Anti-SARS-CoV-2 IgG test (Ortho Vitros test) was performed on an Ortho Clinical Diagnostics Vitros 3600 Immunodiagnostics System at the Mayo Clinic. This assay is approved for clinical testing under FDA Emergency Use Authorization to qualitatively detect antibody to the S1 subunit of SARS-CoV-2 spike protein. Results are reported as reactive (S/CO ≥ 1.0) or nonreactive (S/CO <1.0). Specimens were tested within 7 days of collection and stored at 2-8°C. The same 38 serum samples were run in the Ortho Vitros test, microneutralization assay, and the LFA.

### Lateral Flow Neutralizing Antibody Assay

The Lateral Flow NAb assay was developed to measure levels of antibodies that compete with ACE2 for binding to RBD. The LFA single port cassette (Empowered Diagnostics) contains a test strip composed of a sample pad, blood filter, conjugate pad, nitrocellulose membrane striped with test and control lines, and an absorbent pad (Axim Biotechnologies Inc). The LFA also contains a control mouse antibody conjugated to red gold nanospheres and corresponding anti-mouse IgG striped at the control line.

LFAs were run at room temperature on a flat surface for 10 minutes prior to reading results. To perform the test, 6.7µl of serum or 10ul whole blood were added to the sample port followed by 60µl of chase buffer. After 10 minutes, densities of both test and control lines were recorded in an iDetekt RDS-2500 density reader.

The test leverages the interaction between RBD-conjugated green-gold nanoshells (Nanocomposix) that bind ACE2 at the test line when RBD-neutralizing antibodies (RBD-NAbs) are absent or low. Test line density is inversely proportional to RBD-NAbs present within the sample. As a semi-quantitative test, the results of the LFA can be interpreted using a scorecard or a densitometer. A red line across from the “C” indicates that the test ran properly. An absent or faint test line indicates high levels of RBD-NAbs, whereas a dark test line suggests low or lack of RBD-NAbs.

Precision testing was performed using sera from one highly, and one non-neutralizing donor in replicates of 10. Density values were recorded as above and %CVs calculated using the formula: (Standard Deviation/Mean) * 100%.

### Data Analysis

Pearson’s correlation (*r*) was conducted to assess the strength and significance of associations between the LFA, the Ortho Vitros test and IC50 values. Regression analysis using IC50 values evaluated consistency [14] while Bland-Altman plots assessed agreement and bias [17,18]. Correlation analysis was conducted using IBM SPSS. For two-group analysis, IC50 values corresponding to >240 were categorized as titer of ≥1:320 (neutralizing), whereas IC50 values ≤240 were categorized as ≤1:160 (low/non-neutralizing). Receiver operating characteristic (ROC) analysis was performed to assess accuracy, sensitivity, and specificity of the LFA and Ortho Vitros tests in assessing neutralization; optimal cutoffs for each method were established to maximize area under curve (AUC) [19,20]. ROC analysis was conducted using R language in the RStudio environment (version 3.6.2; RStudio PBC). All analyses were conducted using raw values; data were not normalized, transformed, or scaled.

## RESULTS

As shown at the bottom in **Figure 1A**, serum containing high levels of NAbs results in a weak or ghost test line because NAbs bind RBD on green-gold beads, preventing RBD from binding to the ACE2 receptor at the test line. Serum with low levels of NAbs results in a strong test line because little to no antibodies prevent RBD on beads from binding to ACE2. **Figure 1B** demonstrates results of the test using COVID-19 sera with different levels of NAbs.

**Figure 1.**
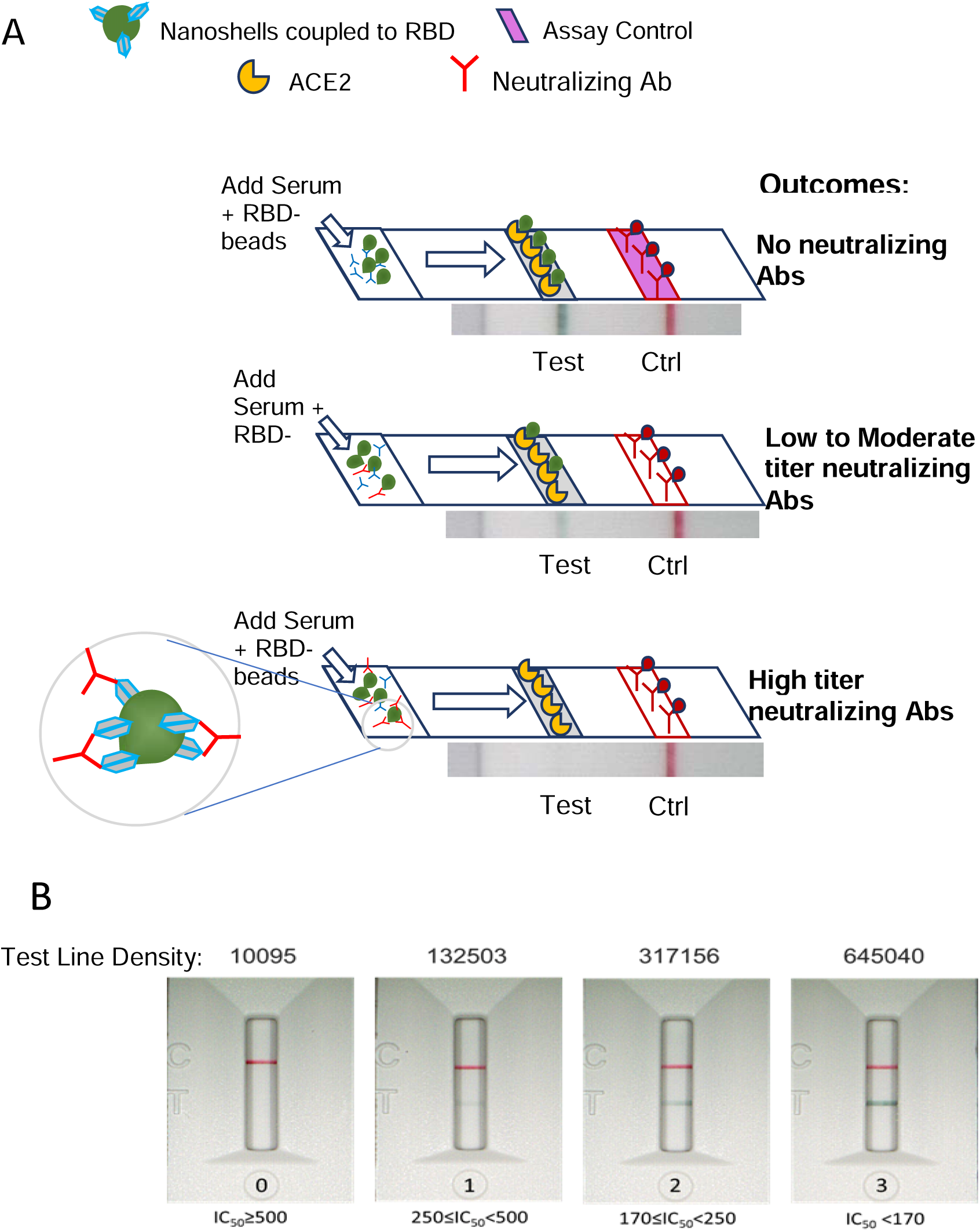
**(A)** Schematic of Neutralization LFA. Below each graphic is a representative image of a lateral flow strip demonstrating actual line density. Addition of non-COVID19-immune serum or plasma (*top*) does not block binding of RBD-beads (green particles) to ACE2 resulting in the RBD-bead–ACE2 complex creating a visible line.

Addition of patient serum with moderate titer NAbs to the sample pad creates a weak line (*middle*). Addition of patient serum with high titer NAbs (> 1:640) blocks binding of RBD-beads to ACE2 such that no line is observed at the test location on the strip (*bottom*). Red control line represents capture of a mouse monoclonal antibody coupled to red beads. **(B)** Scorecard for measuring levels of NAbs. Red control line across from the “C” on the cassette indicates that the test ran properly and the green test line across from the “T” can be used to measure the ability of plasma or serum to block RBD on gold nanoshells from binding to ACE2. **(0)** represents patient serum producing a visually non-existent line with density units of 10,095 and an IC50>500 (IC50=1151); **(1)** represents patient serum with a line density of 132,503 and an IC50 of 396; **(2)** represents patient serum with a line density of 317,156 and an IC50 of 243; (**3)** represents patient serum with a line density of 645,040 and an IC50 of 96.

To support the application of the LFA to measure NAb levels to SARS-CoV-2, we tested 38 serum samples that were assigned IC50 values in a SARS-CoV-2 microneutralization assay [16]. The experiment was performed in a blinded manner such that personnel running either the LFA or the microneutralization assay did not know the results of the comparator test. When line densities from the LFA were plotted against IC50 values determined in the microneutralization assay, serum samples with strong neutralization activity demonstrated low line densities; this indicates that NAbs inhibited RBD from binding to ACE2 (**Figure 2**).

**Figure 2.**
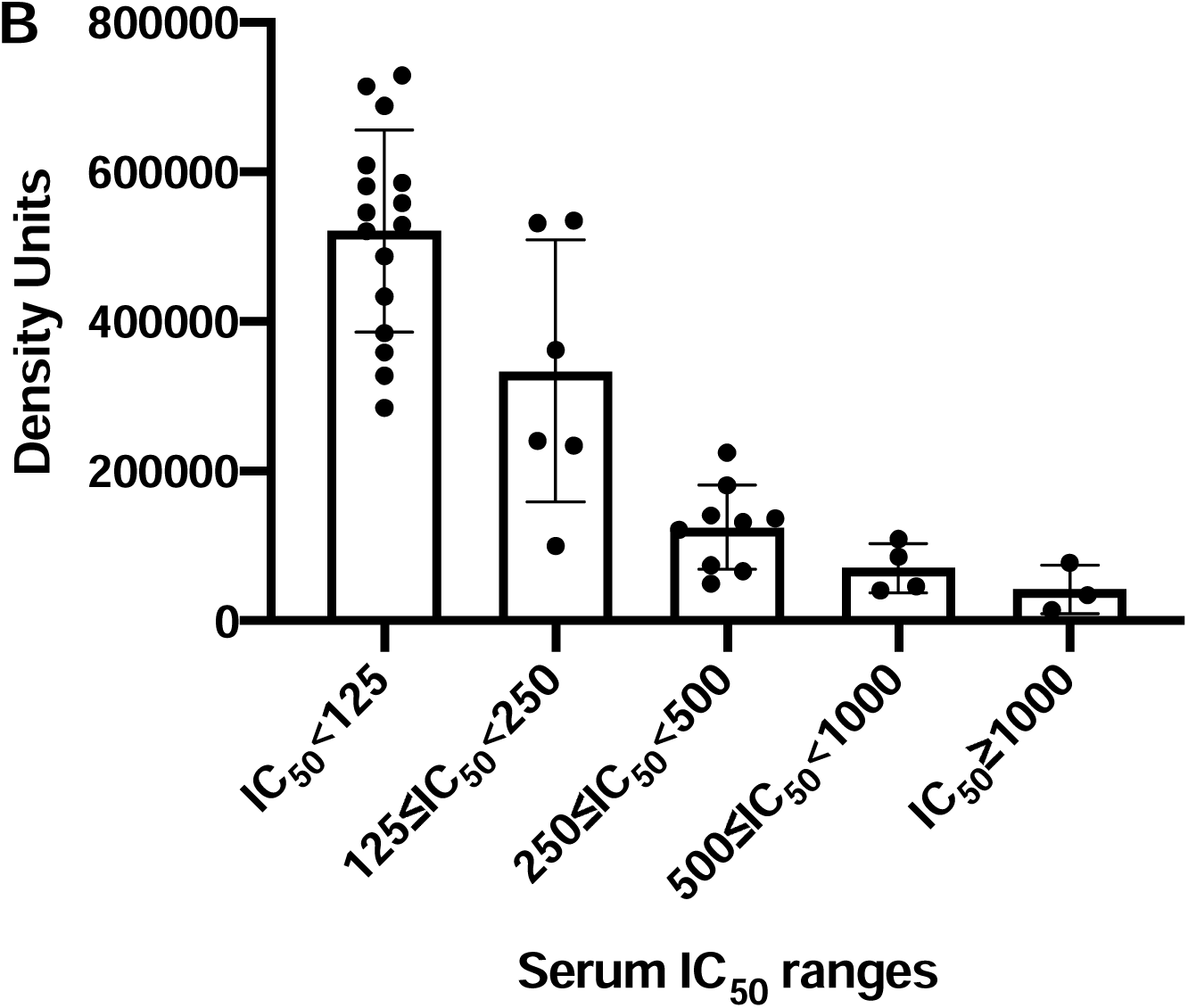
Comparison of RBD-ACE2 competition LFA density values with IC50 values determined in a SARS-CoV-2 microneutralization assay on 38 samples (collected 3 to 90 days after PCR positive result). Ranges of IC50 values are shown on the X-axis plotted against LFA line density units on the Y-axis.

Next, we determined if the LFA detected neutralization activity in serum samples collected from patients with other PCR-confirmed respiratory viruses including seasonal coronaviruses (**Figure 3A**) and for serum samples collected prior to December 2019 (**Figure 3B)**. Neither seasonal respiratory virus sera, nor pre-December 2019 samples showed neutralizing activity.

**Figure 3.**
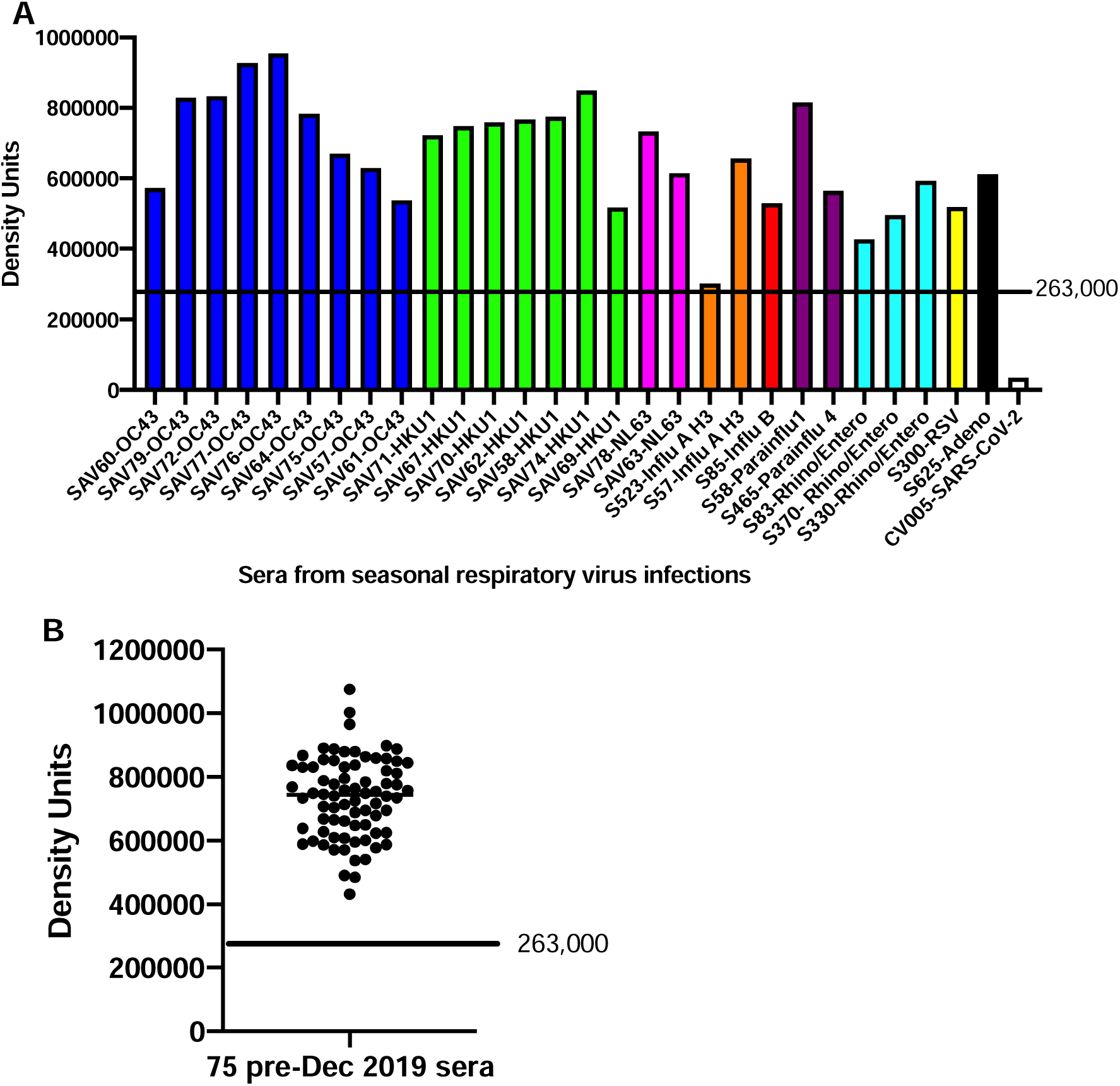
**A)** Serum samples collected with PCR-confirmed diagnosis of seasonal respiratory viruses (Coronavirus OC43, blue; Coronavirus HKU-1, green; Coronavirus NL-63, pink; influenza A, orange, influenza B, red; parainfluenza, purple; rhinovirus, teal; respiratory syncycial virus, yellow; and adenovirus, black were run on the LFA as described in Methods. A positive control serum from a convalescent COVID-19 patient is shown on the far right of the bar graph in white. **B)** Serum samples collected pre-December 2019. Cutoff value of 263,000 density units was calculated based on receiver operating characteristic curves (see Figure 6).

We then compared both the Ortho Vitros test and our LFA to sera with IC50 values determined in the SARS-CoV-2 microneutralization assay using 38 COVID-19 sera. To assess agreement between our LFA and the Ortho Vitros test, density units from the LFA and values from the Ortho test were regressed onto IC50 values (**Supplemental Figure S1**).

**Supplemental Figure S1.**
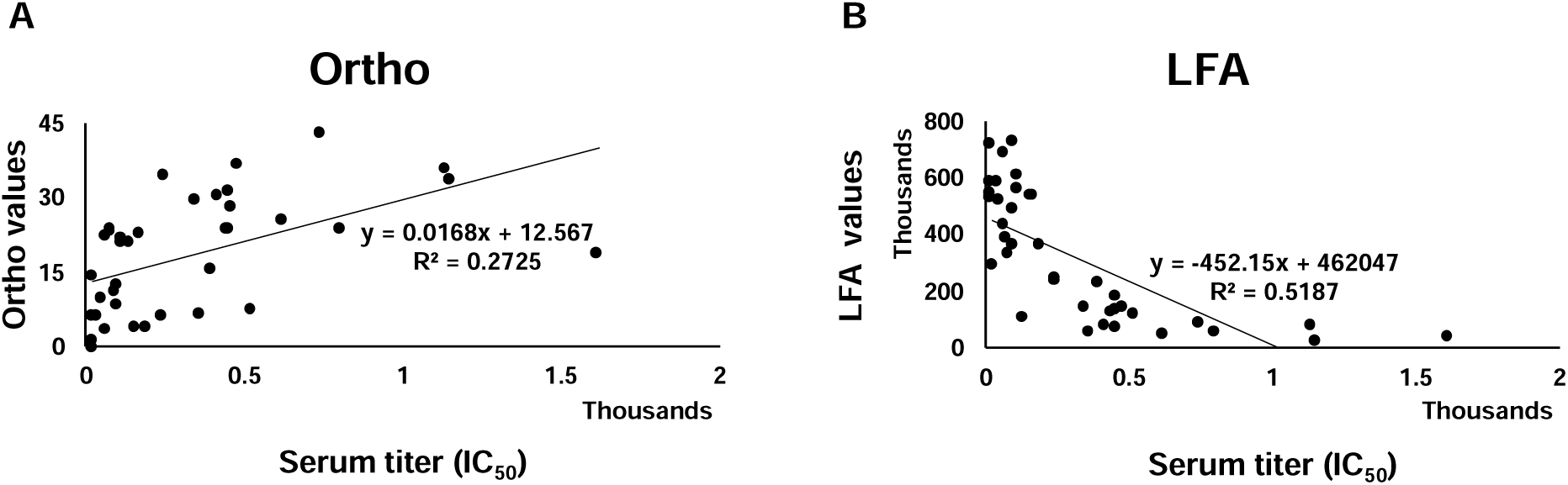
Regression analysis between **(A)** Ortho Vitros SARS-CoV-2 IgG test and **(B)** LFA and titer. Regression plots show explained variance (*R*^*2*^) between compared methods. Thirty-eight samples were tested.

LFA values accounted for roughly 52% of observed variance in IC50 values, while the Ortho Vitros test accounted for approximately 27% of IC50 variance. LFA showed significant negative correlation with IC50 values (*r* = −0.720, *p* < 0.001), while the Ortho Vitros test values showed a significant positive correlation to IC50 values (*r* = 0.522, *p* = 0.001). Additionally, the LFA and Ortho Vitros test values correlated with each other (*r* = −0.572, *p* < 0.001).

To evaluate bias, mean differences and 95% confidence intervals (CIs) were calculated and plotted alongside limits of agreement (**Figure 4**). Both LFA and Ortho Vitros test values showed strong agreement with titer, although the Ortho Vitros test showed a tendency to underestimate neutralizing capacity while the LFA method showed no bias.

**Figure 4.**
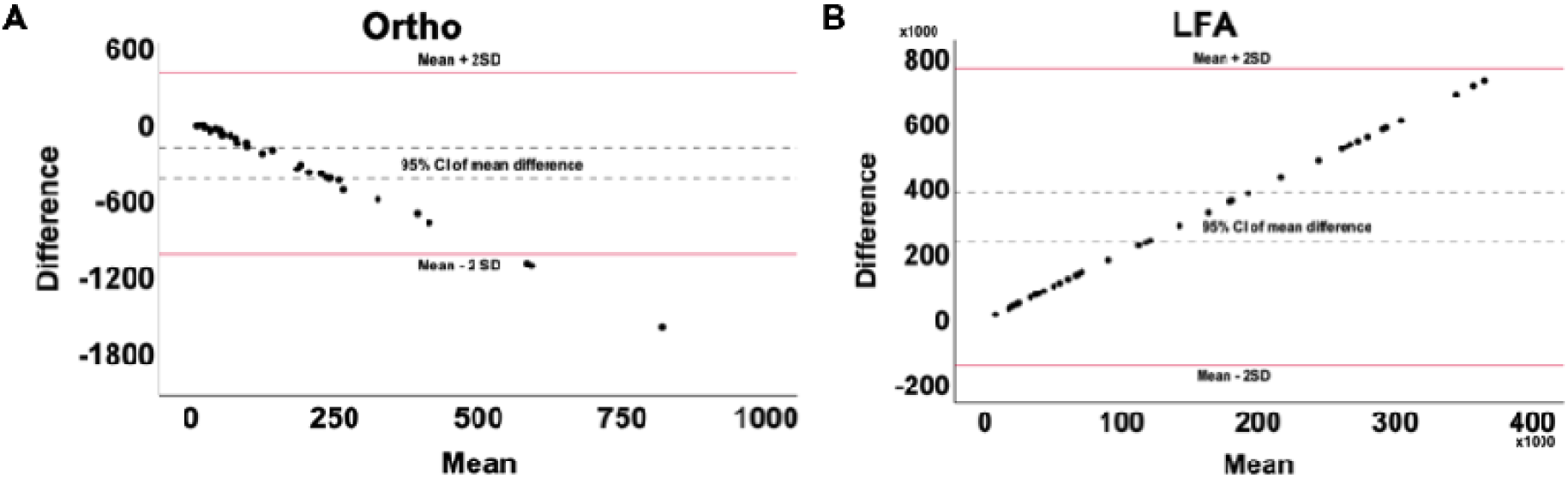
Bland-Altman plots showing bias (mean difference and 95% CI) and computed limits of agreement (mean difference ± 2SD) between **(A)** Ortho Vitros Anti-SARS-CoV-2 IgG test and IC50 values and **(B)** our LFA and IC50 values. Thirty-eight samples were tested.

ROC analysis was performed to assess the ability of the LFA and the Ortho Vitros test to classify low/non-neutralizing (Neg, <1:160), and highly neutralizing groups (≥1:320) (**Figure 5**). As shown in **Figure 5B** and **5D**, the LFA misclassified one non-neutralizing sample (Neg, <1:160) as neutralizing (≥1:320) which the Ortho Vitros test also misclassified as neutralizing. The Ortho test also incorrectly classified five additional neutralizing samples as non-neutralizing.

**Figure 5.**
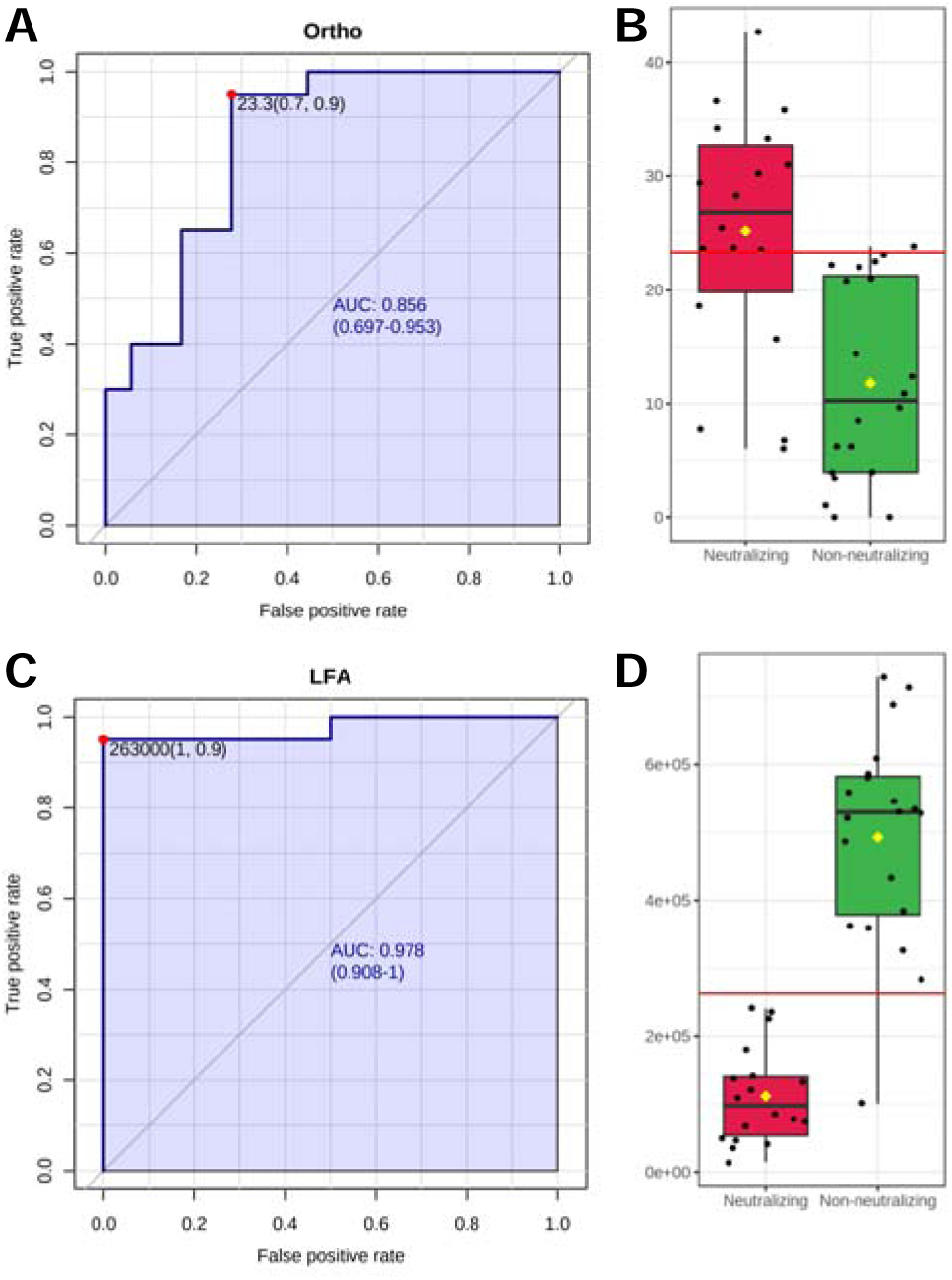
**(A)** Univariate ROC analysis of Ortho Vitros Anti-SARS-CoV-2 IgG test for discrimination of neutralizing samples (≥1:320) [AUC: 0.856, 95% CI: 0.697—0.953, sensitivity = 0.9, specificity = 0.7]. **(B)** Box plot of Ortho Vitros Anti-SARS-CoV-2 IgG test values between neutralizing (≥1:320) and non-neutralizing (Neg—1:160) groups. **(C)** Univariate ROC analysis of LFA for discrimination of neutralizing samples (≥1:320) [AUC: 0.978, 95% CI: 0.908—1.0, sensitivity = 0.9, specificity = 1.0]. **(D)** Box plot of LFA values between neutralizing (≥1:320) and non-neutralizing (Neg—1:160) groups.

Our LFA showed high accuracy for classification of neutralizing samples (AUC = 0.978), while the Ortho Vitros test showed modest accuracy (AUC = 0.856). Notably, while both methods showed roughly 90% sensitivity, the Ortho Vitros test showed only 70% specificity. In contrast, the LFA showed perfect specificity (100%) in this analysis of 38 samples.

Optimal cutoffs were computed to maximize AUC. For the LFA, density unit values below 263,000 classify samples as neutralizing and correspond to titers ≥1:320. Density values above this LFA cutoff classify samples in the non-neutralizing group. For the Ortho Vitros test, values between 0 and 23.3 were representative of non-neutralizing capacity, whereas values above 23.3 were reflective of the neutralizing group.

Precision studies were performed on replicate samples (n=10) and showed a CV of ∼9% from a serum sample in the high neutralizing range and ∼6% CV in a serum sample from the low neutralizing range (Supplemental Table 1).

**Supplemental Table 1.**
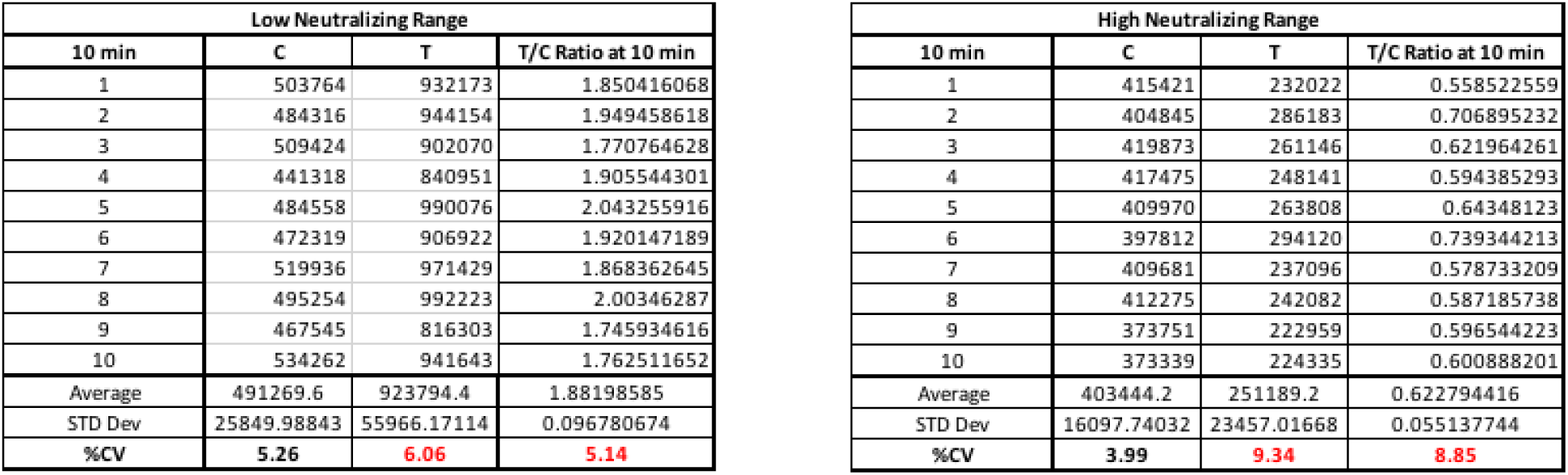
Precision study using one Low Neutralizing Range serum sample and one High Neutralizing Range serum sample in replicates of ten. Low range neutralization is defined as densities from 370,000 – 800,000. The used for precision analysis was from an individual who recovered from COVID-19 but did not neutralize virus in the microneutralization assay (IC50 < 20). High neutralization range samples are defined as densities from 10,000 – 369,999. This sample has an IC50 of 248.

Precision studies were performed on replicate samples (n=10) and showed a CV of ∼9% from a serum sample in the high neutralizing range and ∼6% CV in a serum sample from the low neutralizing range (**Supplemental Table 1**).

Since NAb levels may be considered correlates of protection, we tested sera from RNA vaccine recipients (mRNA-1723 and BNT162b2) in “previously infected” and “not previously infected” individuals using finger-stick blood in the rapid LFA (**Figure 6**). In *previously infected* individuals at baseline (within 3 months of PCR-based diagnosis), 38% demonstrated high levels of NAbs. After the first vaccine dose, 87% of *previously infected* individuals demonstrated high NAb levels, while only 24% of *not previously infected* individuals developed high levels of NAbs. After the second vaccine dose, levels of NAbs increased to 95% in the *previously infected* cohort, while NAb levels increased to 87% in the *not previously infected* cohort. This data suggests that a second vaccine dose is important for highest levels of NAbs.

**Figure 6.**
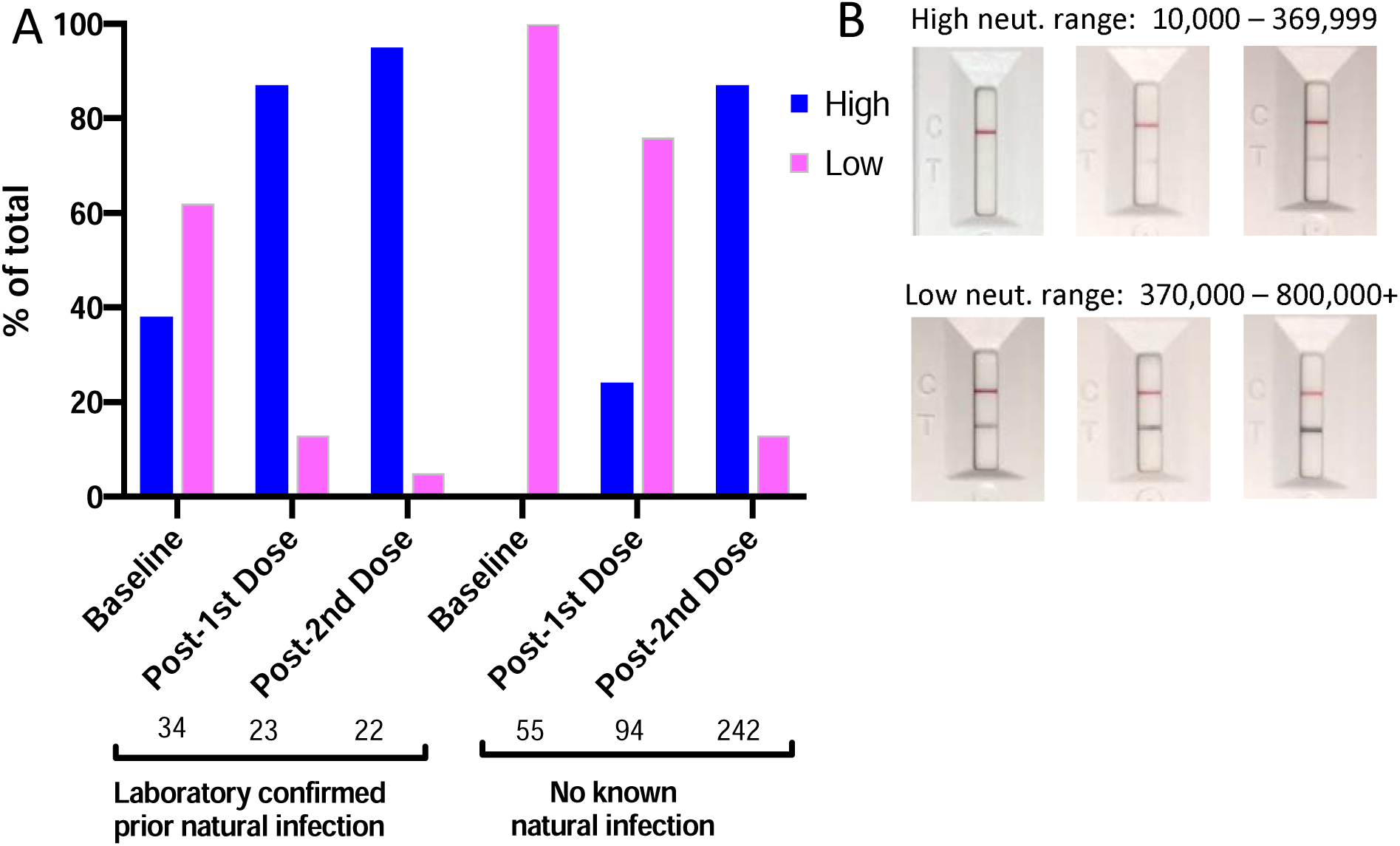
NAb levels in prior infection and vaccine-induced individuals. (A) Baseline indicates within one week of first vaccine dose; Post-1^st^ Dose indicates withing one week of 2^nd^ vaccine dose; Post 2^nd^ Dose indicates 10-20 days after 2^nd^ vaccine dose. High and Low indicates density ranges of Test lines shown in (B). Densities were read in a reader as described in Methods. Serum titers that correspond to high range densities are >1:1280 to ≥1:160. Serum titers corresponding to low range densities are <1:160.

## DISCUSSION

We developed a rapid test that measures levels of NAbs in serum and whole blood. As shown in **Figure 2**, the LFA correlates well with serologic titers determined using a SARS-CoV-2 microneutralization assay, especially when serum sample IC50 values are >250. Advantages of the LFA test are that it can be inexpensively and rapidly deployed to determine levels of NAbs in vaccine recipients. Moreover, the test can be used longitudinally to evaluate duration of protective immunity in naturally infected and vaccinated individuals–many more than could ever be evaluated using BSL2 or BSL3-based neutralization assays.

The LFA and Ortho Vitros test showed a significant correlation with each other (*r* = −0.572, *p* < 0.001), displaying good linear relation (r = −0.720, p < 0.001)[21]. The LFA accounts for 52% of observed IC50 variance (*R*^*2*^ = 0.5187), while the Ortho Vitros test accounts for 27% (*R*^*2*^ = 0.2725). Although absolute quantitation demands an excellent coefficient of determination (*R*^*2*^ ≥ 0.99)[22], variables with *R*^*2*^ ≥ 0.5 are highly predictive in univariate regression models while measures with *R*^*2*^ < 0.5 are recommended for use in multivariate models with complementary measures to increase predictive accuracy [23,24]. Bland-Altman analysis (**Figure 4**) showed the Ortho Vitros test to be prone to underestimation of IC50 values, while the LFA method did not exhibit over- or underestimation bias. Furthermore, across mean values for both methods, the LFA showed discrete differential values while the Ortho Vitros test struggled to differentiate high neutralizing samples.

Using our rapid test to measure NAbs in previously infected vaccine recipients and those who were not infected agrees with other studies in BSL3 facilities using serum from venipuncture blood [5,25–29]. Natural infection may not elicit high levels of NAbs [6–8], but a first dose of vaccine induces high levels of NAbs in the majority of recipients similar to 2 doses of vaccine in non-previously infected individuals, suggesting natural infection primes the immune system[30]. In naïve individuals, a single dose of vaccine elicits high NAb levels (Titers >1:160) in only 24% of vaccine recipients, leaving 76% of vaccine recipients with titers lower than 1:160 which would not qualify for convalescent plasma donation according to FDA memo of March 9,2021. After a second vaccine dose, the LFA indicated high levels of NAbs in 87% of recipients, identical to levels observed in previously infected individuals after the first vaccine dose. These findings might suggest that a booster (3^rd^ vaccine dose) in non-infected individuals could induce the highest levels of NAbs in the most people.

Limitations of the LFA are that it uses only the RBD portion of spike protein. Although the vast majority of reports indicate that the principle neutralizing domain is the RBD portion of spike protein, mAbs have been reported that neutralize SARS-CoV-2 by binding to the N-terminal domain of spike protein [31,32]. Also, since the spike protein assumes multiple conformations during viral binding and entry [33], neutralizing epitopes exist on the quaternary structure of spike [32]. Although RBDs on the nanoparticles may associate, it is not known if they assume a native conformation.

Other limitations are the binary nature of this data analysis (high and low neutralizing) of a continuous assay. NAb levels should be evaluated longitudinally to assess rise and fall in NAb levels; this rapid test is well-suited for that role. Another limitation is that the LFA does not differentiate high affinity anti-RBD NAbs from an abundance of lower affinity anti-RBD NAbs.

This test may prove useful in monitoring COVID-19 vaccine recipients as a correlate of protection. It would be logistically difficult to obtain a tube of blood from every vaccine recipient for BSL3 work. However, since this LFA requires only a drop of blood, individual use of this test might lead to more comprehensive longitudinal monitoring of protective humoral immunity and indicate when boosters might be required.

## Data Availability

All data in the manuscript is available.

## AUTHOR CONTRIBUTIONS

DL, SS and AS-N developed the LFA. AJ performed the experiments and interpreted the results. PJ performed statistical analysis. EK, TG and KM contributed samples. CK, AJ and KP tested vaccine recipients and interpreted results. SP, NK and AB performed SARS-CoV-2 microneutralization assays and interpreted results LW and JM contributed samples for preliminary experiments. MM-G designed the strip layout and produced the strips for the vaccine study.

## CONFLICTS OF INTEREST STATEMENT

DL and SS are co-founders of Sapphire, the research division of Axim Biotech. SS, MM-G and AS-N are employees of Axim Biotech. Other authors have no conflicts regarding this work.

## FUNDING

This study was funded in part by Axim Biotech, San Diego, CA

## Notes

### Competing Interest Statement

AJR, EK, PJ, SP, NK, AB, TG, LW, JRM and KM have no competing interests. DFL received support from Axim Biotechnologies to perform the study. SS and AS-N are supported and employed by Axim Biotechnologies.

### Funding Statement

Axim Biotechnologies supported this work.

### Author Declarations

Peripheral blood and serum were collected for this study under an Arizona State University IRB approved protocol #0601000548 and Mayo Clinic IRB protocol #20-004544.

### Summary of Updates

Supplementary Figure legend S1 incorrectly referred to S1A as S1B. It is now correct.

